# Predictive performance of multi-model ensemble forecasts of COVID-19 across European nations

**DOI:** 10.1101/2022.06.16.22276024

**Authors:** K. Sherratt, H. Gruson, R. Grah, H. Johnson, R. Niehus, B. Prasse, F. Sandman, J. Deuschel, D. Wolffram, S. Abbott, A. Ullrich, G. Gibson, EL. Ray, NG. Reich, D. Sheldon, Y. Wang, N. Wattanachit, L. Wang, J. Trnka, G. Obozinski, T. Sun, D. Thanou, L. Pottier, E. Krymova, MV. Barbarossa, N. Leithäuser, J. Mohring, J. Schneider, J. Wlazlo, J. Fuhrmann, B. Lange, I. Rodiah, P. Baccam, H. Gurung, S. Stage, B. Suchoski, J. Budzinski, R. Walraven, I. Villanueva, V. Tucek, M. Šmíd, M. Zajícek, C. Pérez Álvarez, B. Reina, NI. Bosse, S. Meakin, P. Alaimo Di Loro, A. Maruotti, V. Eclerová, A. Kraus, D. Kraus, L. Pribylova, B. Dimitris, ML. Li, S. Saksham, J. Dehning, S. Mohr, V. Priesemann, G. Redlarski, B. Bejar, G. Ardenghi, N. Parolini, G. Ziarelli, W. Bock, S. Heyder, T. Hotz, D. E. Singh, M. Guzman-Merino, JL. Aznarte, D. Moriña, S. Alonso, E. Álvarez, D. López, C. Prats, JP. Burgard, A. Rodloff, T. Zimmermann, A. Kuhlmann, J. Zibert, F. Pennoni, F. Divino, M. Català, G. Lovison, P. Giudici, B. Tarantino, F. Bartolucci, G. Jona Lasinio, M. Mingione, A. Farcomeni, A. Srivastava, P. Montero-Manso, A. Adiga, B. Hurt, B. Lewis, M. Marathe, P. Porebski, S. Venkatramanan, R. Bartczuk, F. Dreger, A. Gambin, K. Gogolewski, M. Gruziel-Slomka, B. Krupa, A. Moszynski, K. Niedzielewski, J. Nowosielski, M. Radwan, F. Rakowski, M. Semeniuk, E. Szczurek, J. Zielinski, J. Kisielewski, B. Pabjan, K. Holger, Y. Kheifetz, M. Scholz, M. Bodych, M. Filinski, R. Idzikowski, T. Krueger, T. Ozanski, J. Bracher, S. Funk

## Abstract

**Background:** Short-term forecasts of infectious disease burden can contribute to situational awareness and aid capacity planning. Based on best practice in other fields and recent insights in infectious disease epidemiology, one can maximise the predictive performance of such forecasts if multiple models are combined into an ensemble. Here we report on the performance of ensembles in predicting COVID-19 cases and deaths across Europe between 08 March 2021 and 07 March 2022.

**Methods:** We used open-source tools to develop a public European COVID-19 Forecast Hub. We invited groups globally to contribute weekly forecasts for COVID-19 cases and deaths reported from a standardised source over the next one to four weeks. Teams submitted forecasts from March 2021 using standardised quantiles of the predictive distribution. Each week we created an ensemble forecast, where each predictive quantile was calculated as the equally-weighted average (initially the mean and then from 26th July the median) of all individual models’ predictive quantiles. We measured the performance of each model using the relative Weighted Interval Score (WIS), comparing models’ forecast accuracy relative to all other models. We retrospectively explored alternative methods for ensemble forecasts, including weighted averages based on models’ past predictive performance.

**Results:** Over 52 weeks we collected and combined up to 28 forecast models for 32 countries. We found a weekly ensemble had a consistently strong performance across countries over time. Across all horizons and locations, the ensemble performed better on relative WIS than 84% of participating models’ forecasts of incident cases (with a total N=862), and 92% of participating models’ forecasts of deaths (N=746). Across a one to four week time horizon, ensemble performance declined with longer forecast periods when forecasting cases, but remained stable over four weeks for incident death forecasts. In every forecast across 32 countries, the ensemble outperformed most contributing models when forecasting either cases or deaths, frequently outperforming all of its individual component models. Among several choices of ensemble methods we found that the most influential and best choice was to use a median average of models instead of using the mean, regardless of methods of weighting component forecast models.

**Conclusions:** Our results support the use of combining forecasts from individual models into an ensemble in order to improve predictive performance across epidemiological targets and populations during infectious disease epidemics. Our findings further suggest that median ensemble methods yield better predictive performance more than ones based on means. Our findings also highlight that forecast consumers should place more weight on incident death forecasts than incident case forecasts at forecast horizons greater than two weeks.

**Code and data availability:** All data and code are publicly available on Github: covid19-forecast-hub-europe/euro-hub-ensemble.

## Background

Epidemiological forecasts make quantitative statements about a disease outcome in the near future. Forecasting targets can include measures of prevalent or incident disease and its severity, for some population over a specified time horizon. Researchers, policy makers, and the general public have used such forecasts to understand and respond to the global outbreaks of COVID-19 [1]–[3].

Forecasters use a variety of methods and models for creating and publishing forecasts, varying in both defining the forecast outcome and in reporting the probability distribution of outcomes [4], [5]. Such variation makes it difficult to compare predictive performance between forecast models, and from there to derive objective arguments for using one forecast over another. This confounds the selection of a single representative forecast and reduces the reliability of the evidence base for decisions based on forecasts.

A “forecast hub” is a centralised effort to improve the transparency and usefulness of forecasts, by standardising and collating the work of many independent teams producing forecasts [6]. A hub sets a commonly agreed-upon structure for forecast targets, such as type of disease event, spatio-temporal units, or the set of quantiles of the probability distribution to include from probabilistic forecasts. For instance, a hub may collect predictions of the total number of cases reported in a given country for each day in the next two weeks. Forecasters can adopt this format and contribute forecasts for centralised storage in the public domain. This shared infrastructure allows forecasts produced from diverse teams and methods to be visualised and quantitatively compared on a like-for-like basis, which can strengthen public and policy use of disease forecasts.

The underlying approach to creating a forecast hub was pioneered in climate modelling and adapted for collaborative epidemiological forecasts of dengue [7] and influenza in the USA [6], [8]. This infrastructure was adapted for forecasts of short-term COVID-19 cases and deaths in the US [9], [10], prompting similar efforts in some European countries [11]–[13].

Standardising forecasts allows for combining multiple forecasts into a single ensemble with the potential for an improved predictive performance. Evidence from previous efforts in multi-model infectious disease forecasting suggests that forecasts from an ensemble of models can be consistently high performing compared to any one of the component models [7], [8], [14]. Elsewhere, weather forecasting has a long-standing use of building ensembles of models using diverse methods with standardised data and formatting in order to improve performance [15], [16].

The European COVID-19 Forecast Hub [17] is a project to collate short term forecasts of COVID-19 across 32 countries in the European region. The Hub is funded and supported by the European Centre for Disease Prevention and Control (ECDC), with the primary aim to provide reliable information about the near-term epidemiology of the COVID-19 pandemic to the research and policy communities and the general public [3]. Second, the Hub aims to create infrastructure for storing and analysing epidemiological forecasts made in real time by diverse research teams and methods across Europe. Third, the Hub aims to maintain a community of infectious disease modellers underpinned by open science principles.

We started formally collating and combining contributions to the European Forecast Hub in March 2021. Here, we investigate the predictive performance of an ensemble of all forecasts contributed to the Hub in real time each week, as well as the performance of variations of ensemble methods created retrospectively.

## Methods

We developed infrastructure to host and analyse prospective forecasts of COVID-19 cases and deaths. The infrastructure is compatible with equivalent research software from the US [18], [19] and German and Polish COVID-19 [20] Forecast Hubs, and easy to replicate for new forecasting collaborations.

### Forecast targets and models

We sought forecasts for the incidence of COVID-19 as the total reported number of cases and deaths per week. We considered forecasts for 32 countries in Europe, including all countries of the European Union, European Free Trade Area, and the United Kingdom. We compared forecasts against observed data reported for each country by Johns Hopkins University (JHU, [21]). JHU data sources included a mix of national and aggregated subnational data. We aggregated incidence over the Morbidity and Mortality Weekly Report (MMWR) epidemiological week definition of Sunday through Saturday.

Teams could express their uncertainty around any single forecast target by submitting predictions for up to 23 quantiles (from 0.01 to 0.99) of the predictive probability distribution. Teams could also submit a single point forecast. At the first submission we asked teams to add a pre-specified set of metadata briefly describing the forecasting team and methods (see supplementary information (SI)). No restrictions were placed on who could submit forecasts. To increase participation we actively contacted known forecasting teams across Europe and the US and advertised among the ECDC network. Teams submitted a broad spectrum of model types, ranging from mechanistic to empirical models, agent-based and statistical models, and ensembles of multiple quantitative or qualitative models (described at [22]). We maintain a full project specification with a detailed submissions protocol [23].

We collected forecasts submitted weekly in real time over the 52 week period from 08 March 2021 to 07 March 2022. Teams submitted at latest two days after the complete dataset for the latest forecasting week became available each Sunday. We implemented an automated validation programme to check that each new forecast conformed to standardised formatting. Forecast validation ensured a monotonic increase of predictions with each increasing quantile, integer-valued non-negative counts of predicted cases, as well as consistent date and location definitions.

Each week we used all available valid forecasts to create a weekly real-time ensemble model (referred to as “the ensemble” from here on), for each of the 256 possible forecast targets: incident cases and deaths in 32 locations over the following one through four weeks. The ensemble method was an unweighted average of all models’ forecast values, at each predictive quantile for a given location, target, and horizon. From 08 March 2021, we used the arithmetic mean. However we noticed that including highly anomalous forecasts in a mean ensemble produced extremely wide uncertainty. To mitigate this, from 26^th^ July 2021 onwards the ensemble instead used a median of all predictive quantiles.

We created an open and publicly accessible interface to the forecasts and ensemble, including an online visualisation tool allowing viewers to see past data and interact with one or multiple forecasts for each country and target for up to four weeks’ horizon [24]. All forecast and meta data are freely available and held on Github [17] and Zoltar, a platform for hosting epidemiological forecasts [25], [26]. In the codebase for this study [27] we provide a simple method and instructions for downloading and preparing these data for analysis using R. We encourage other researchers to freely use and adapt this to support their own analyses.

### Forecast evaluation

In this study we focus only on the comparative performance of forecasting models relative to each other. For each model, we evaluated performance in terms of both accuracy (coverage) and overall predictive performance (weighted interval score). We evaluated all previous forecasts against actual observed values for each model, stratified by the forecast horizon, location, and target. We calculated scores using the *scoringutils* R package [28]. We removed any forecast surrounding (both the week of, and the first week after) a strongly anomalous data point. We defined anomalous as where any subsequent data release revised that data point by over 5%.

We established the accuracy of each model’s prediction boundaries as the coverage of the predictive intervals. We calculated coverage at a given interval level k, where *k* ∈ [0,1], as the proportion *p* of observations that fell within the corresponding central predictive intervals across locations and forecast dates. A perfectly calibrated model would have *p* = *k* at all 11 levels (corresponding to 22 quantiles excluding the median). An underconfident model at level *k* would have *p > k*, i.e. more observations fall within a given interval than expected. In contrast, an overconfident model at level *k* would have *p < k*, i.e. fewer observations fall within a given interval than expected. We here focus on coverage at the *k* = 0.5 and *k* = 0.95 levels.

We also assessed the overall predictive performance of weekly forecasts using the weighted interval score (WIS) across all available quantiles. The WIS represents a parsimonious approach to scoring forecasts based on uncertainty represented as forecast values across a set of quantiles [29], and is a strictly proper scoring rule, that is, it is optimal for predictions that come from the data-generating model. As a consequence, the WIS encourages forecasters to report predictions representing their true belief about the future [30]. Each forecast for a given location and date is scored based on an observed count of weekly incidence, the median of the predictive distribution and the predictive upper and lower quantiles corresponding to the central predictive interval level.

Not all models provided forecasts for all locations and dates, and we needed to compare predictive performance in the face of various levels of missingness across each forecast target. Therefore we calculated a relative WIS. This is a measure of forecast performance which takes into account that different teams may not cover the same set of forecast targets (i.e., weeks and locations). The relative WIS is computed using a *pairwise comparison tournament* where for each pair of models a mean score ratio is computed based on the set of shared targets. The relative WIS of a model with respect to another model is then the ratio of their respective geometric mean of the mean score ratios, such that smaller values indicate better performance.

We scaled the relative WIS of each model with the relative WIS of a baseline model, for each forecast target, location, date, and horizon. The baseline model assumes case or death counts stay the same as the latest data point over all future horizons, with expanding uncertainty, described previously in [31]. Here we report the relative WIS of each model with respect to the baseline model.

### Retrospective ensemble methods

We retrospectively explored alternative methods for combining forecasts for each target at each week. A natural way to combine probability distributions available in the quantile format used here is [32]

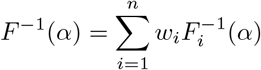

Where *F*_1_ … *F*_*n*_ are the cumulative distribution functions of the individual probability distributions (in our case, the predictive distributions of each forecast model *i* contributed to the hub), *w*_*i*_ are a set of weights in [0,1]; and *α* are the quantile levels such that

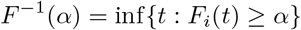

Different ensemble choices then mainly translate to the choice of weights *w*_*i*_. An arithmetic mean ensemble uses weights at *w*_*i*_ = 1*/n*, where all weights are equal and sum up to 1.

Alternatively, we can choose a set of weights to apply to forecasts before they are combined. Numerous options exist for choosing these weights with the aim to maximise predictive performance, including choosing weights to reflect each forecast’s past performance (thereby moving from an untrained to a trained ensemble). A straightforward choice is so-called inverse score weighting. In this case, the weights are calculated as

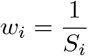

where *S*_*i*_ reflects the forecasting skill of forecaster *i*, normalised so that weights sum to 1. This method of weighting was found in the US to outperform unweighted scores during some time periods [33] but this was not confirmed in a similar study in Germany and Poland [11].

When constructing ensembles from quantile means, a single outlier can have an oversized effect on the ensemble forecast. Previous research has found that a median ensemble, replacing the arithmetic mean of each quantile with a median of the same values, yields competitive performance while maintaining robustness to outlying forecasts [34]. Building on this, we also created weighted median ensembles using the weights described above and a Harrel-Davis quantile estimator with a beta function to approximate the weighted percentiles [35]. We then compared the performance of unweighted and inverse relative WIS weighted mean and median ensembles.

## Results

An example of weekly forecasts from the ensemble model is shown in Figure 1.

**Figure 1:**
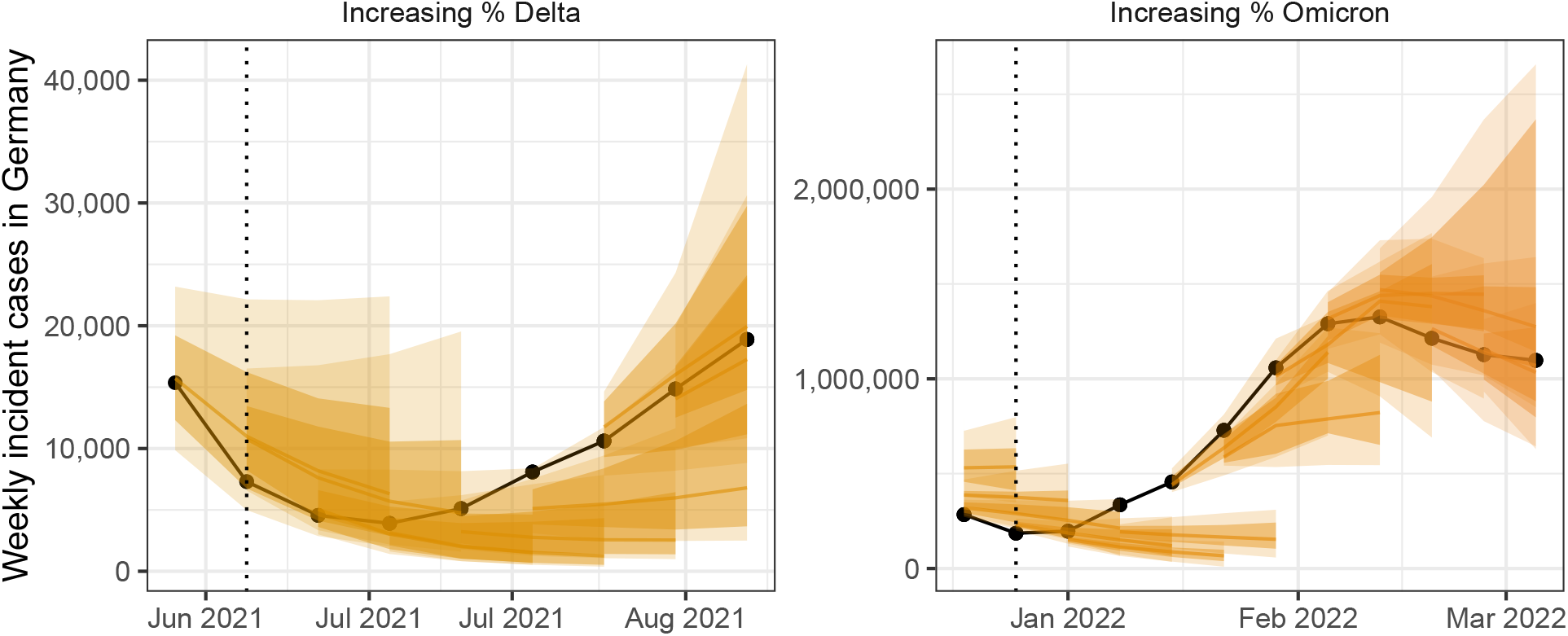
Ensemble forecasts of weekly incident cases in Germany over periods of increasing SARS-CoV-2 variants Delta (B.1.617.2, left) and Omicron (B.1.1.529, right). Black indicates observed data. Coloured ribbons represent each weekly forecast of 1-4 weeks ahead (showing median, 50%, and 90% probability). For each variant, forecasts are shown over an x-axis bounded by the earliest dates at which 5% and 99% of sequenced cases were identified as the respective variant of concern, while vertical dotted lines indicate the approximate date that the variant reached dominance (>50% sequenced cases).

Over the whole study period, 26 independently participating forecasting teams contributed results from 28 unique forecasting models (see supplementary information (SI), Table 1). The number of models contributing to each ensemble forecast varied over time and by forecasting target (SI Figure 1). Not all modellers created forecasts for all locations, horizons, or variables. At most, 15 models contributed forecasts for cases in Germany at the 1 week horizon, with an accumulated 592 forecasts for that single target over the study period. In contrast, deaths in Finland at the 2 week horizon saw the smallest number of forecasts, with only 6 independent models contributing a total 24 forecasts. Similarly, not all teams forecast across all quantiles of the predictive distribution for each target, with only 23 models providing the full set of 23 quantiles. No ensemble forecast was composed of less than 3 independent models.

**Table 1:** Predictive performance of main ensembles, as measured by the scaled relative WIS.

| Horizon | Weighted mean | Weighted median | Unweighted mean | Unweighted median |
| --- | --- | --- | --- | --- |
| **Cases** |  |  |  |  |
| 1 week | 0.59 | 0.62 | 0.59 | 0.61 |
| 2 weeks | 0.67 | 0.67 | 0.67 | 0.67 |
| 3 weeks | 0.79 | 0.70 | 0.81 | 0.71 |
| 4 weeks | 1.06 | 0.75 | 1.09 | 0.79 |
| **Deaths** |  |  |  |  |
| 1 week | 0.63 | 0.59 | 1.00 | 0.59 |
| 2 weeks | 0.57 | 0.54 | 0.81 | 0.53 |
| 3 weeks | 0.64 | 0.56 | 0.83 | 0.54 |
| 4 weeks | 0.83 | 0.64 | 0.82 | 0.62 |

Using all models and the ensemble, we created 2106 forecasting scores where each score summarises a unique combination of forecasting model, variable, country, and week ahead horizon (SI Figure 2). We visually compared the absolute performance of forecasts in predicting numbers of incident cases and deaths. We observed that forecasts predicted well in times of stable epidemic behaviour, while struggling to accurately predict at longer horizons around inflection points, for example during rapid changes in population-level behaviour or surveillance. Forecast models varied widely in their ability to predict and account for the introduction of new variants, giving the ensemble forecast over these periods a high level of uncertainty (Figure 1).

In relative terms, the ensemble of all models performed well compared to both its component models and the baseline. By relative WIS scaled against a baseline of 1 (where a score <1 indicates outperforming the baseline), the median score for participating models across all submitted forecasts was 1.04, while the median score of forecasts from the ensemble model was 0.71. Across all horizons and locations, the ensemble performed better on scaled relative WIS than 84% of participating model scores when forecasting cases (with a total N=862), and 92% of participating model scores for forecasts of incident deaths (N=746).

The performance of individual and ensemble forecasts varied by length of the forecast horizon (Figure 2). At each horizon, the typical performance of the ensemble outperformed both the baseline model and the aggregated scores of all its component models, although we saw wide variation between individual models in performance across horizons. Both individual models and the ensemble saw a trend of worsening performance at longer horizons when forecasting cases with the median scaled relative WIS of the ensemble across locations worsened from 0.62 for one-week ahead forecasts to 0.9 when forecasting four weeks ahead. Performance for forecasts of deaths was more stable over one through four weeks, with median ensemble performance moving from 0.69 to 0.76 across the four week horizons.

**Figure 2:**
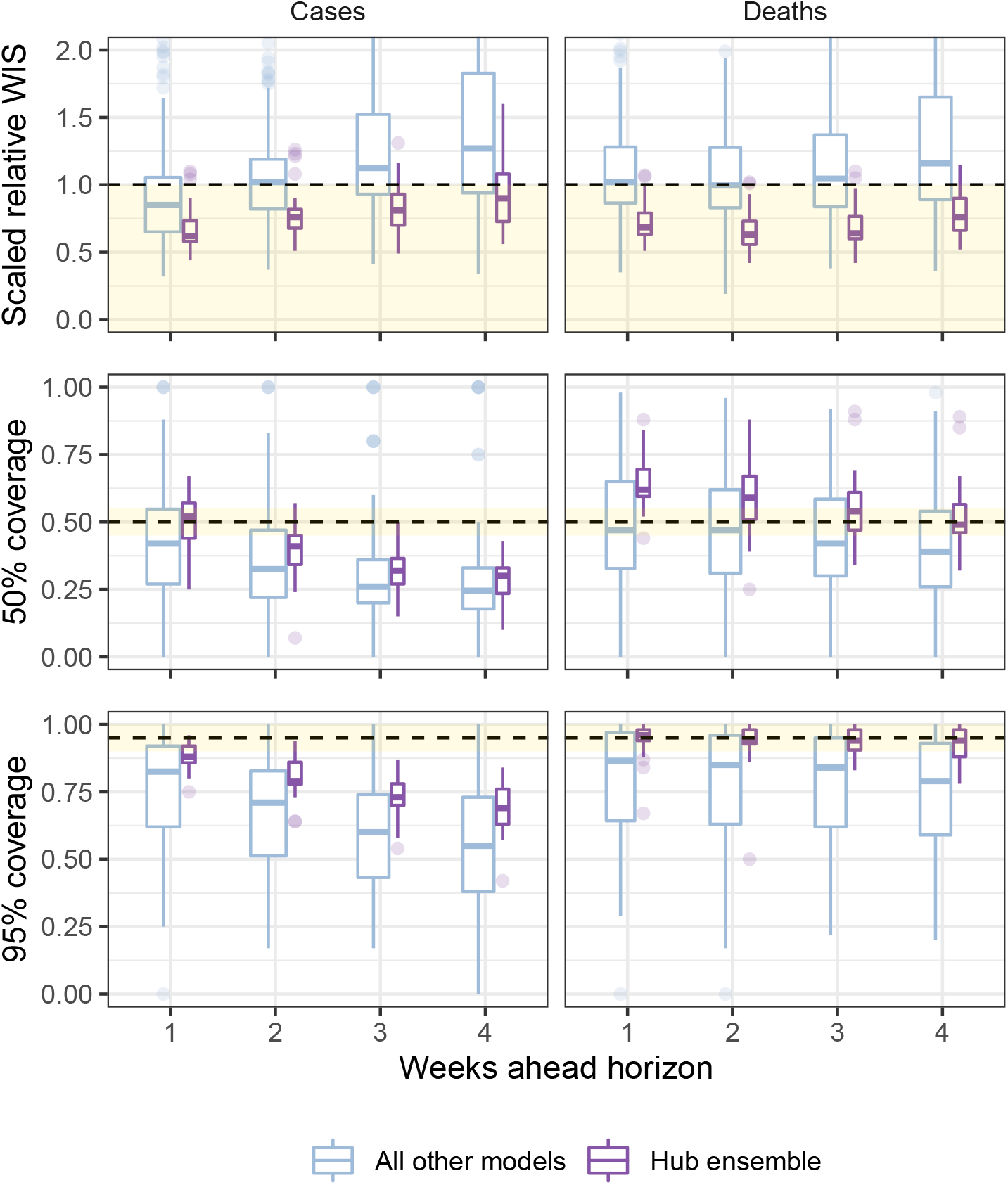
Performance of short-term forecasts aggregated across all individually submitted models and the Hub ensemble, by horizon, forecasting cases (left) and deaths (right). Performance measured by relative weighted interval score scaled against a baseline (dotted line, 1), and coverage of uncertainty at the 50% and 95% levels. Boxplot, with width proportional to number of observations, show interquartile ranges with outlying scores as faded points. The target range for each set of scores is shaded in yellow.

We observed similar trends in performance across horizon when considering how well the ensemble was calibrated with respect to the observed data. At one week ahead the case ensemble was well calibrated (ca. 50% and 95% nominal coverage at the 50% and 95% levels respectively). This did not hold at longer forecast horizons as the case forecasts became increasingly over-confident. Meanwhile, the ensemble of death forecasts was well calibrated at the 95% level across all horizons, and the calibration of death forecasts at the 50% level improved with lengthening horizons compared to being underconfident at shorter horizons.

The ensemble also performed consistently well in comparison to individual models when forecasting across countries (Figure 3). In total, across 32 countries forecasting for one through four weeks, when forecasting cases the ensemble outperformed 75% of component models in 21 countries, and outperformed all available models in 3 countries. When forecasting deaths, the ensemble outperformed 75% and 100% of models in 30 and 9 countries respectively. Considering only the the two-week horizon shown in Figure 3, the ensemble of case forecasts outperformed 75% models in 24 countries and all models in only 12 countries. At the two-week horizon for forecasts of deaths, the ensemble outperformed 75% and 100% of its component models in 30 and 26 countries respectively.

**Figure 3:**
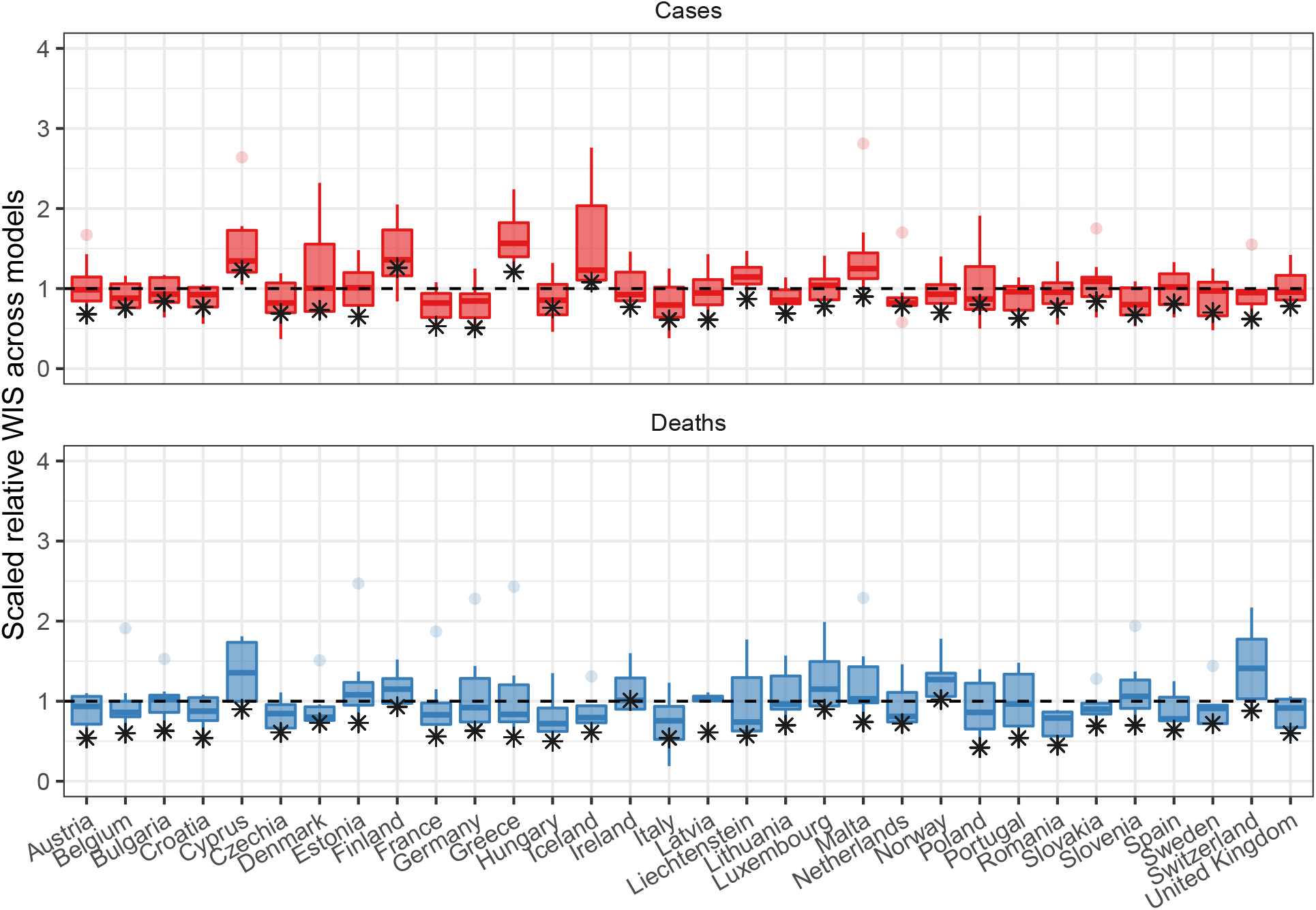
Performance of short-term forecasts across models and median ensemble (asterisk), by country, forecasting cases (top) and deaths (bottom) for two-week ahead forecasts, according to the relative weighted interval score. Boxplots show interquartile ranges, with outliers as faded points, and the ensemble model performance is marked by an asterisk. y-axis is cut-off to an upper bound of 4 for readability.

We considered alternative methods for creating ensembles from the participating forecasts, using either a mean or median to combine either weighted or unweighted forecasts (Table 1). Across locations we observed that the median outperformed the mean across all one through four week horizons and both cases and death targets, for all but cases at the 1 week horizon. This held regardless of whether the component forecasts were weighted or unweighted by their individual past performance. Between methods of combination, weighting made little difference to the performance of the median ensemble, but slightly improved performance of the mean ensemble.

## Discussion

We collated 12 months of forecasts of COVID-19 cases and deaths across 32 countries in Europe, collecting from multiple independent teams and using a principled approach to standardising both forecast targets and the predictive distribution of forecasts. We combined these into an ensemble forecast and compared the relative performance of forecasts between models, finding that the ensemble forecasts outperformed most individual models across all countries and horizons over time.

Across all models we observed that forecasting changes in trend in real time was particularly challenging. Our study period included multiple fundamental changes in viral-, individual-, and population-level factors driving the transmission of COVID-19 across Europe. In early 2021, the introduction of vaccination started to change population-level associations between infections, cases, and deaths [36], while the Delta variant emerged and became dominant [37]. Similarly from late 2021 we saw the interaction of individually waning immunity during the emergence and global spread of the Omicron variant [38]. Neither the extent nor timing of these factors were uniform across European countries covered by the Forecast Hub [39]. This meant that the performance of any single forecasting model depended partly on the ability, speed, and precision with which it could adapt to new conditions for each forecast target.

We observed a contrast between a more stable performance of forecasting deaths further into the future compared to forecasts of cases. Previous work has found rapidly declining performance for case forecasts with increasing horizon [31], [40], while death forecasts can perform well with up to six weeks lead time [41]. We can similarly link this to the specific epidemic dynamics in this study. COVID-19 has a typical serial interval of less than a week [42]. This implies that case forecasts of more than two weeks only remain valid if rates of transmission and detection remain stable over the entire forecast horizon. This is unlikely to have held given the rapid changes in epidemic dynamics across many countries in Europe. Meanwhile, we can interpret the higher reliability of death forecasts as due to the longer time lag between infection and death [43], and higher consistency of reporting deaths in surveillance data [44]. This allows forecasters to incorporate the effect of changes in transmission. Additionally, the performance of trend-based forecasts may have benefited from the slower changes to trends in incident deaths caused by gradually increasing vaccination rates.

We found the ensemble in this study continued to outperform both other models and the baseline at up to four weeks ahead. Our results support previous findings that ensemble forecasts are the best or nearly the best performing models with respect to absolute predictive performance and appropriate coverage of uncertainty [12], [14], [31]. While the ensemble was consistently high performing, it was not strictly dominant across all forecast targets, reflecting findings from previous comparable studies of COVID-19 forecasts [11], [45]. Our finding suggests the usefulness of an ensemble as a robust summary when forecasting across many spatiotemporal targets, without replacing the importance of communicating the full range of model predictions.

When exploring variations in ensemble methods, we found that the choice of median over means yielded the most consistent improvement in predictive performance, regardless of the method of weighting. Other work has supported the importance of the median in providing a stable forecast that better accounts for outlier forecasts than the mean [45], although though this finding may be dependent on the quality of the individual forecast submissions. In contrast, weighing models by past performance did not result in any consistent improvement in performance. This is in line with existing mixed evidence for any optimal ensemble method for combining short term probabilistic infectious disease forecasts. Many methods of combination have performed competitively in analyses of forecasts for COVID-19 in the US, including the simple mean and weighted approaches outperforming unweighted or median methods [33]. This contrasts with later analyses finding weighted methods to give similar performance to a median average [10], [45]. We can partly explain this inconsistency if performance of each method depends on the outcome being predicted (cases, deaths), its count (incident, cumulative) and absolute level, the changing disease dynamics, and the varying quality and quantity of forecasting teams over time.

We note several limitations in our approach to assessing the relative performance of an ensemble among forecast models. Our results are the outcome of evaluating forecasts against a specific performance metric and baseline, where multiple options for evaluation exist and the choice reflects the aim of the evaluation process. Further, our choice of baseline model affects the given performance scores in absolute terms, and more generally the choice of appropriate baseline for epidemic forecast models is not obvious when assessing infectious disease forecasts. The model used here is supported by previous work [31], yet previous evaluation in a similar context has suggested that choice of baseline affects relative performance in general [46], and future research should be done on the best choices of baseline models in the context of infectious disease epidemics.

Our assessment of forecast performance may further have been inaccurate due to limitations in the observed data against which we evaluated forecasts. We sourced data from a globally aggregated database to maintain compatibility across 32 countries [21]. However, this made it difficult to identify the origin of lags and inconsistencies between national data streams, and to what extent these could bias forecasts for different targets. In particular we saw some real time data revised retrospectively, introducing bias in either direction where the data used to create forecasts was not the same as that used to evaluate it. We attempted to mitigate this by using an automated process for determining data revisions, and excluding forecasts made at a time of missing, unreliable, or heavily revised data. More generally it is unclear if the expectation of observation revisions should be a feature built into forecasts. Further research is needed to understand the perspective of end-users of forecasts in order to assess this.

In an emergency setting, open access to visualised forecasts and underlying data is useful for researchers, policymakers, and the public. For forecast producers, an easily accessible comparison between results from different methods can highlight individual strengths and weaknesses and help prioritise new areas of work. For forecast users, probabilistic information about the future can influence decisions in the present that can then change epidemic dynamics [1].

Existing participatory modelling efforts for COVID-19 have been useful for policy communication [2], while multi-country efforts have included only single models adapted to country-specific parameters [47]–[49]. By expanding participation to many modelling teams, our work was able to create robust ensemble forecasts across Europe while allowing comparison across forecasts built with different interpretations of current data, on a like for like scale in real time. At the same time, collating time-stamped predictions ensures that we can test true out-of-sample performance of models and avoid retrospective claims of performance. Testing the limits of forecasting ability with these comparisons forms an important part of communicating any model-based prediction to decision makers.

This study raises many further questions which could inform epidemic forecast modellers and users. The dataset created by the European Forecast Hub is an openly accessible, standardised, and extensively documented catalogue of real time forecasting work from a range of teams and models across Europe [24], and we recommend its use for further research on forecast performance. In the code developed for this study we provide a worked example of downloading and using both the forecasts and their evaluation scores [27].

Future work could explore the impact on forecast models of changing epidemiology at a broad spatial scale by combining analyses of trends and turning points in cases and deaths with forecast performance, or extending to include data on vaccination, variant, or policy changes over time. There is also much scope for future research into methods for combining forecasts to improve performance of an ensemble. This includes altering the inclusion criteria of forecast models based on different thresholds of past performance, excluding or including only forecasts that predict the lowest and highest values (trimming) [33], or using alternative weighting methods such as quantile regression averaging [12]. Exploring these questions would add to our understanding of real time performance, supporting and improving future forecasting efforts.

We see additional scope to adapt the Hub format to the changing COVID-19 situation across Europe. We have extended the Forecast Hub infrastructure to include short term forecasts for hospitalisations with COVID-19, which is a challenging task due to limited data across the locations covered by the hub. As the policy focus shifts from immediate response to anticipating changes brought by vaccinations or the geographic spread of new variants [39], we are also separately investigating models for longer term scenarios in addition to the short term forecasts in a similar framework to existing scenario modelling work in the US [50].

In conclusion, we have shown that during a rapidly evolving epidemic spreading through multiple populations, an ensemble forecast performed highly consistently across a large matrix of forecast targets, typically outperforming the majority of its separate component models and a naive baseline model. In addition, we have linked issues with the predictability of short-term case forecasts to underlying COVID-19 epidemiology, and shown that ensemble methods based on past model performance were unable to reliably improve forecast performance. Our work constitutes a step towards both unifying COVID-19 forecasts and improving our understanding of them.

## Data Availability

All source data were openly available before the study, originally available at: https://github.com/covid19-forecast-hub-europe/covid19-forecast-hub-europe. All data and code for this study are openly available on Github: covid19-forecast-hub-europe/euro-hub-ensemble.

https://github.com/covid19-forecast-hub-europe/euro-hub-ensemble

https://github.com/covid19-forecast-hub-europe/covid19-forecast-hub-europe

