## Supplement: SI table 1, SI figure 1, SI figure 2 for "Predictive performance of multi-model ensemble forecasts of COVID-19 across European nations"

### Participating teams

The following teams participated in the European Forecast Hub by contributing forecasts over the study period. Information below is taken from metadata provided by each team.

| Model | Affiliation | Methods | Metadata |
|---|---|---|---|
| BIOCOMSC-Gompertz | BIOCOMSC | Empirical model based on cases and deaths dynamics. | Metadata |
| CovidMetrics-epiBATS | University of Cologne Covid Metrics | Forecasts are based on TBATS - models (DeLivera, Hyndman and Snyder (2011)) and are updated daily for each German state. | Metadata |
| epiforecasts-EpiNow2 | Epiforecasts / London School of Hygiene and Tropical Medicine | Semi-mechanistic estimation of the time-varying reproduction number for latent infections mapped to reported cases/deaths. | Metadata |
| epiforecasts-weeklygrowth | epiforecasts | A Bayesian autoregressive model using weekly incidence data, application of the forecast.vocs R package. | Metadata |
| epiMOX-SUIHTER | epiMOX | Compartmental model SUIHTER | Metadata |
| EuroCOVIDhub-ensemble | European COVID-19 Forecast Hub | An ensemble, or model average, of submitted forecasts to the European COVID-19 Forecast Hub. | Metadata |
| FIAS_FZJ-Epi1Ger | Frankfurt Institute for Advanced Studies & Forschungszentrum Jülich | An extended SEIR model with additional compartments for undetected cases | Metadata |

| Model | Affiliation | Methods | Metadata |
|---|---|---|---|
| HZI-AgeExtendedSEIR | Helmholtz Zentrum fuer Infektionsforschung | Deterministic SEIR type model | Metadata |
| ICM-agentModel | ICM / University of Warsaw | Agent-based model | Metadata |
| IEM_Health-CovidProject | IEM Health | SEIR model projections for daily incident confirmed COVID cases and deaths by using AI to fit actual cases observed. | Metadata |
| ILM-EKF | ILM | Extended Kalman filter based on reproduction equation | Metadata |
| itwm-dSEIR | Fraunhofer Institute for Industrial Mathematics ITWM | cohort based, integral equation | Metadata |
| ITWW-county_repro | ITWW | Forecasts of county level incidence based on regional reproduction numbers. | Metadata |
| JBUD-HMXK | JBUD | Heavily modified infection-age SIR-X model with waning immunity, vaccinations, seasonality and undetected cases. | Metadata |
| MOCOS-agent1 | MOCOS group | Agent-based microsimulation model | Metadata |
| MUNI-ARIMA | Masaryk University | ARIMA model with outlier detection fitted to transformed weekly aggregated series. | Metadata |
| MUNI_DMS-SEIAR | Department of Mathematics and Statistics Masaryk University Team | SEIAR model with A compartment of absent unobserved infected estimated from hospital data with incorporated mobility data dependence; optimized to the compartment of all exposed (unobserved included) | Metadata |
| PL_GRedlarski-DistrictsSum | Grzegorz Redlarski | Modified SIR method, applied to all districts. Forecasts for districts are summed up. | Metadata |

| Model | Affiliation | Methods | Metadata |
|---|---|---|---|
| prolix-euclidean | prolix | Offsets obtained by correlations, best linear approximation of reproduction rates (using vaccination approximation) by least euclidean distance, and linear prediction. | Metadata |
| RobertWalraven-ESG | Robert Walraven | Multiple skewed gaussian distribution peaks fit to raw data | Metadata |
| SDSC_ISG-TrendModel | Swiss Data Science Center / University of Geneva | The Trend Model predicts daily cases and deaths using linear extrapolation on the linear or log scale of the underlying trend estimated by a robust LOESS seasonal-trend decomposition model. | Metadata |
| Statgroup19-richards | Statgroup19 | Richards' curve based generalized growth model | Metadata |
| Statgroup19-spatialrichards | Statgroup19 | Richards' curve based generalized growth model taking into account spatial dependence | Metadata |
| UC3M-EpiGraph | Universidad Carlos III de Madrid | Agent-based parallel simulator that models individual interactions extracted from social networks and demographical data. | Metadata |
| ULZF-SEIRC19SI | University of Ljubljana, Faculty of Health Sciences Team | SEIHR model extended with compartments for hospitals, intensive care units, asymptomatic cases, separate submodels for vaccinated and unvaccinated, divided to 5 age subgroups of population | Metadata |
| UMass-MechBayes | UMass-Amherst | Bayesian compartmental model with observations on cumulative case counts and cumulative deaths. Model is fit independently to each state. Model includes observation noise and a case detection rate. | Metadata |
| UpgUmibUsi-MultiBayes | UNIPG_UNIMIB_USI_UNINSUBRIA | Bayesian Dirichlet-Multinomial models for counts of patients in mutually exclusive and exhaustive categories such as hospitalized in regular wards and in intensive care units, deceased and recovered | Metadata |

| Model | Affiliation | Methods | Metadata |
| --- | --- | --- | --- |
| USC-SIkJalpha | University of Southern California | A heterogeneous infection rate model with human mobility for epidemic modeling. Our model adapts to changing trends and provide predictions of confirmed cases and deaths. | Metadata |
| UVA-Ensemble | University of Virginia, Biocomplexity COVID-19 Response Team | An ensemble of multiple methods such as auto-regressive (AR)models with exogenous variables, Long short-term memory (lSTM) models,Kalman filter and PatchSim (an SEIR model). | Metadata |

**Summary of evaluated forecasts**

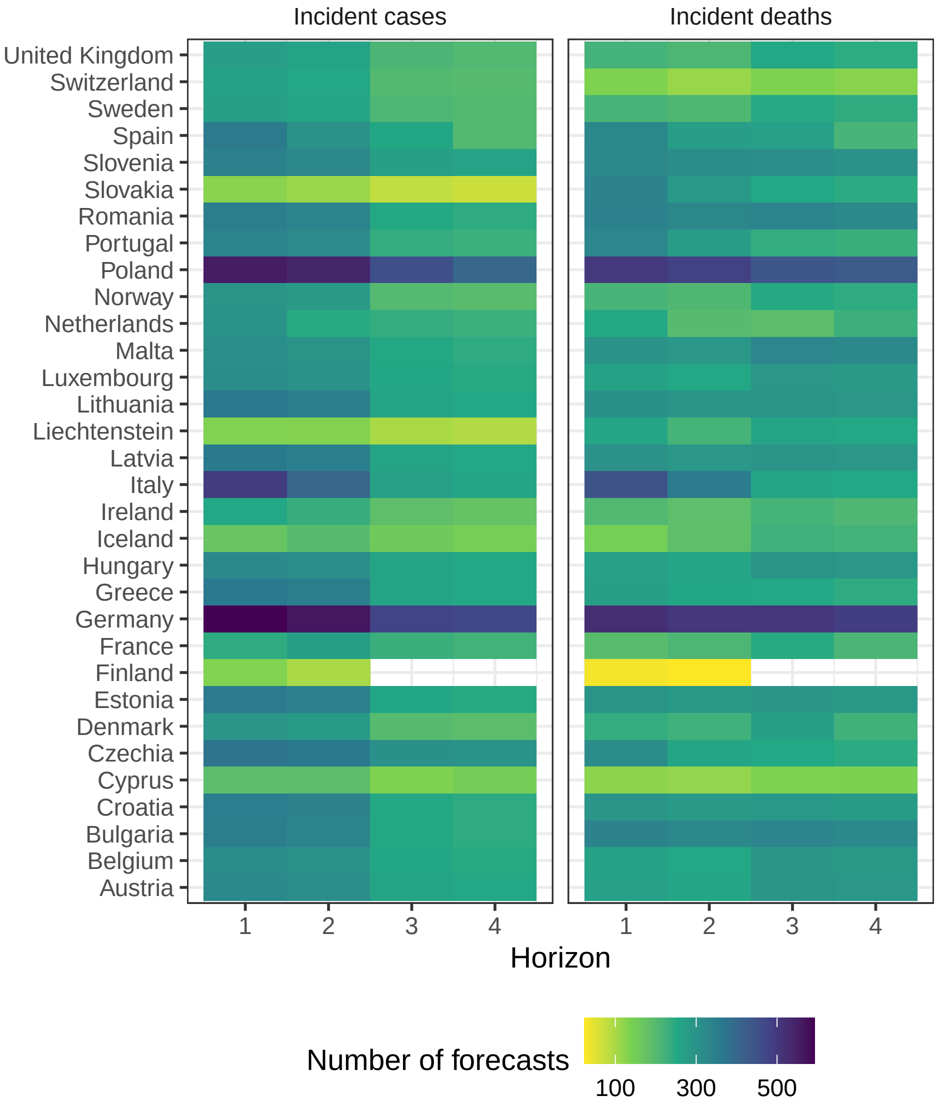


Figure 1: Total number of forecasts included in evaluation, by target location, week ahead horizon, and variable

**Comparison of contributed forecasts and the Hub ensemble**

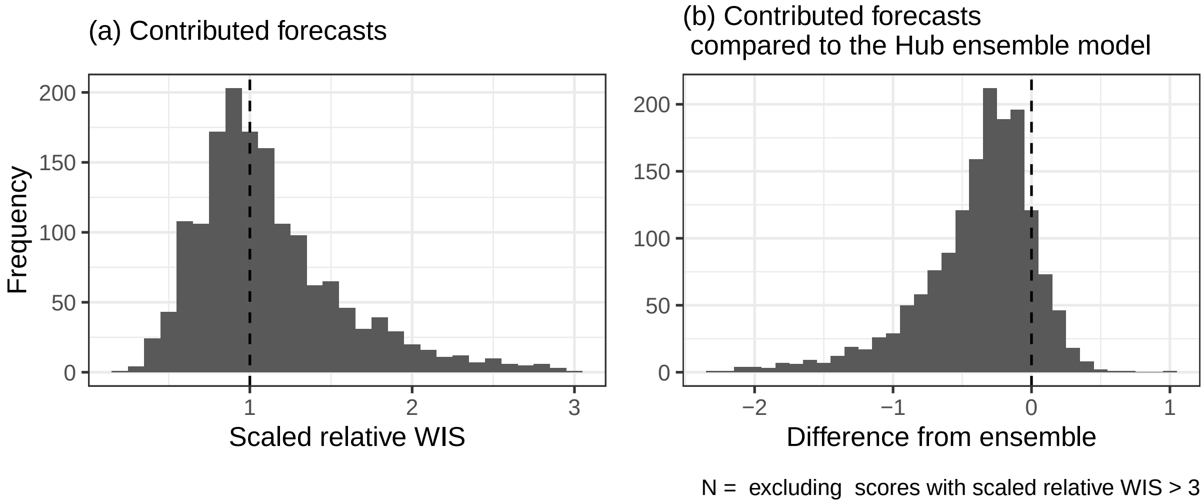


Figure 2: Comparison of scores between participating model forecasts and Hub ensemble of all available forecasts for each target
